# X- vs. Y-Chromosome Influences on Human Behavior: A Deep Phenotypic Comparison of Psychopathology in XXY and XYY Syndromes

**DOI:** 10.1101/2023.06.19.23291614

**Authors:** Lukas Schaffer, Srishti Rau, Liv Clasen, Allysa Warling, Ethan T. Whitman, Ajay Nadig, Cassidy McDermott, Anastasia Xenophontos, Kathleen Wilson, Jonathan Blumenthal, Erin Torres, Armin Raznahan

**Affiliations:** Section on Developmental Neurogenomics, Human Genetics Branch, National Institute of Mental Health Intramural Research Program, Bethesda, MD, USA; Center for Autism Spectrum Disorders and Division of Neuropsychology, Children’s National Hospital, Washington DC, USA

## Abstract

Do different genetic disorders impart different psychiatric risk profiles? This question has major implications for biological and translational aspects of psychiatry, but has been difficult to tackle given limited access to shared batteries of fine-grained clinical data across genetic disorders. Using a new suite of generalizable analytic approaches, we examine gold-standard diagnostic ratings, scores on 66 dimensional measures of psychopathology, and measures of cognition and functioning in two different sex chromosome aneuploidies (SCAs) – Klinefelter (XXY/KS) and XYY syndrome (n=102 and 64 vs. n=74 and 60 matched XY controls, total n=300). We focus on SCAs for their high collective prevalence, informativeness regarding differential X– vs. Y-chromosome effects, and potential relevance for normative sex differences. We show that XXY/KS elevates rates for most psychiatric diagnoses as previously reported for XYY, but disproportionately so for anxiety disorders. Fine-mapping across all 66 traits provides a detailed profile of psychopathology in XXY/KS which is strongly correlated with that of XYY (r=.75 across traits) and robust to ascertainment biases, but reveals: (i) a greater penetrance of XYY than KS/XXY for most traits except mood/anxiety problems, and (ii) a disproportionate impact of XYY vs. XXY/KS on social problems. XXY/KS and XXY showed a similar coupling of psychopathology with adaptive function and caregiver strain, but not IQ. This work provides new tools for deep-phenotypic comparisons of genetic disorders in psychiatry and uses these to detail unique and shared effects of the X– and Y-chromosome on human behavior.

## INTRODUCTION

Gene dosage disorders (GDDs) – encompassing sub-chromosomal copy number variations and aneuploidies – are well-recognized, highly penetrant neuropsychiatric risk factors [1, 2]. In addition to being important medical conditions in their own right, GDDs also hold broader relevance for biological psychiatry as naturally occurring models of gene dosage effects on human neurodevelopment and behavior [3–6]. For recurrent GDDs (due to errors in chromosomal disjunction and homologous recombination [7]), it is possible to recruit and phenotype cohorts of individuals carrying the same genomic alteration [8, 9]. Genetics-first research approaches in recurrent GDDs offer an objective foothold in the complex biology of neuropsychiatric disorders and a way of mapping genotype-phenotype relationships in humans for mechanistic insights and potential advances in precision psychiatry [5, 10, 11]. However, fully harnessing this potential requires detailed comparison of phenotypic profiles between different GDDs.

There has been growing recent interest in comparative psychiatric phenotyping of GDDs, and available studies suggest that while there is substantial variability in the penetrance of any given GDD, there are also noteworthy differences in the profile of psychiatric risk imparted by different GDDs [12–16].To date however, comparative behavioral studies between different GDDs have generally not used gold-standard interviewer-based diagnostic instruments for all participants [17–19] and have been limited to relatively few dimensions/measures of psychopathology [12, 14]. Furthermore, existing literature has not compared GDDs in the mapping between psychopathology and clinically relevant cognitive and functional features (CFFs) such as general cognitive ability, adaptive functioning, and caregiver strain.

Notably, the representation of different GDDs in comparative studies has generally not scaled with GDD prevalence, meaning that we still lack deep phenotypic comparisons for the most common family of human GDDs: sex chromosome aneuploidies (SCAs, collective prevalence ∼ 1:500 live births [20, 21]). Most published comparisons of SCAs have been limited in their focus on only a few concurrent phenotypes (e.g., eight syndrome scales from the Child Behavior Checklist [15]) or phenotypes which fall within a restricted behavioral domain (e.g., social deficits [22, 23], ADHD symptoms [24, 25], or language abilities [26, 27], or multidimensional measures in modest sample sizes lacking diagnostic information [28].

To address these gaps in knowledge, we present a deep-phenotypic comparison between two of the most common SCAs in males – XXY (Klinefelter syndrome, henceforth XXY/KS, estimated prevalence ∼ 1:650 live male births [20, 21] and XYY syndrome (estimated prevalence ∼ 1:1000 live male births [20, 21]. This work is part of an ongoing prospective natural history study of SCAs at the NIMH Intramural Research Program where different SCA groups are characterized using the same detailed assessment battery. Our study design incorporates gold-standard diagnostic interviews, multi-dimensional measures of psychopathology, estimates of the coupling between psychopathology and CFFs, and tests for evidence of ascertainment bias by comparing pre– and postnatally diagnosed SCA subgroups. We recently reported findings in XYY using this study design [29]. The current report applies the same detailed characterization to XXY/KS syndrome and systematically compares XXY/KS and XYY for rates of psychiatric diagnosis, profiles of scores across 66 measured dimensions of psychopathology and the coupling between these dimensions and CFFs.

Our study design is not just important for better understanding XXY/KS and XYY as collectively common neurogenetic syndromes in their own right, but carries broader impact for psychiatric genetics in three key areas. First, sex chromosome influences on the brain could potentially contribute to well-known sex-differences in psychiatric risk within the general population, and SCAs provide models for probing X– and Y– chromosome influences on the human brain ([30, 31]. Second, SCAs benefit from a wealth of preexisting comparative data from transcriptomic and neuroimaging analyses [31–34]. Consequently, psychiatric comparisons of XXY/KS and XYY provide a unique opportunity to assess how the degree of phenotypic convergence vs. divergence for a given pair of GDDs at the level of behavior coheres with that across different levels of biological description (e.g., gene expression, neuroanatomy). Finally, the analytic approaches developed and applied here for comparative phenotyping of SCAs represent a new methodological toolkit that is immediately generalizable for comparative research in any other clinical groups of interest.

## METHODS

### Participant Recruitment

The present study includes data from a total of 300 individuals comprising two case-control cohorts: (i) a new, previously unreported XXY/KS cohort comprising 102 individuals with XXY/KS syndrome and 74 age– and sex-matched typically developing XY controls, and (ii) a previously characterized XYY cohort (64 XYY individuals and 60 XY controls, described in [29]). For both XXY/KS and XYY cohorts the matched XY controls provided reference score distributions for a battery of behavioral and psychiatric measurement scales, against which observed scores in SCA individuals could be normalized. This approach allows scores for all dimensional measures in a diverse questionnaire battery to be expressed on the same continuous z-score scale despite the fact that some rating scales do not provide scaled scores relative to published population norms (see below). As such, the analyses below consider both categorical diagnoses and z-scored dimensional measures of psychopathology for 64 XYY and 102 XXY/KS individuals.

Participants with SCA were recruited in collaboration with the Association for X and Y Chromosome Variations (AXYS; genetic.org) and the National Institutes of Health (NIH) Clinical Center Office of Patient Recruitment (which also assisted in the recruitment of XY controls). All XY controls were screened using a standardized interview to verify the absence of a prior psychiatric diagnosis or early developmental difficulties requiring provision of extra support at school or home. The sole inclusion criteria for XYY and XXY/KS individuals in this study was cytogenetic confirmation of their SCA karyotype (non-mosaic based on 50 metaphase spreads), and age between 5 and 25 years on the day of assessment. For SCA participants who were able to give blood (n=37 XYY, n=99 XXY/KS) the SCA was re-confirmed through repeat karyotype testing (Quest Diagnostics, Nichols Institute, Chantilly, VA). For the remaining participants (n=27 XYY, n=3 XXY/KS), karyotypic eligibility was verified by inspection of existing community-based genetic testing reports. Exclusion criteria for both the XXY/KS and XY control participants included a history of brain injury or comorbid neurological disorders. All research assessments were conducted at the NIH Clinical Center, Bethesda, MD, USA. Written informed consent was secured from adult participants and parents of minor participants and written assent from children. The study was approved by the National Institute of Mental Health (NIMH) Institutional Review Board.

### Participant Characterization

#### Clinical History

All participants received a structured medical history and physical examination. Caregiver reports and reviews of prior medical documents were used to record the age of SCA diagnosis. Caregiver strain was measured using the Caregiver Strain Questionnaire (CSQ) [35].

#### Diagnostic Assessments

The K-SADS-PL (henceforth, “KSADS” [36]) was administered to all SCA participants (with the exception of one XXY/KS individual who could not be interviewed) by a trained psychiatric nurse practitioner (ET), and all final consensus ratings were reviewed with a child and adolescent psychiatrist (AR). The KSADS is a semi-structured instrument used with both the participant and parent/guardian (assessed separately) to diagnose psychiatric disorders based on the Diagnostic and Statistical Manual of Mental Disorders-Fifth Edition (DSM-V). It was used to generate diagnostic prevalence rates. All components of the KSADS were administered in this study, except the autism spectrum disorder (ASD) section. Formal ASD clinical assessment consisted of three components: the Autism Diagnostic Observation Schedule, 2^nd^ Edition (ADOS [37], the Autism Diagnostic Interview, Revised (ADI-R [38], and a DSM-V ASD diagnostic criteria checklist [39]. Diagnostic classification or ASD were based on ADI/ADOS administration in all SCA individuals except 15/102 XXY/KS individuals for whom these assessments could not be completed. Of these 15, we included 11 in our diagnostic description who had not previously been diagnosed with ASD and had SCQ scores below 7. As such, the denominator for the ASD diagnostic rate in XXY/KS is 98. Assessment were completed by doctorate level clinicians with extensive ASD evaluation experience, who met research reliability standards on the ADI-R and ADOS-2. For descriptive purposes, we also calculated collective diagnostic rates for groups of disorders defined as follows: Neurodevelopmental Disorders (including ASD, ADHD, Tic Disorder, Obsessive Compulsive Disorder); Mood Disorders (including Bipolar Disorder, Major Depressive Disorder, Disruptive Mood Dysregulation Disorder); Anxiety Disorders; Post-traumatic Stress and Related Disorders; Disruptive, Impulse Control and Conduct Disorders (including Oppositional Defiant Disorder and Conduct Disorder); and Substance Use Disorders.

#### Battery of Questionnaire-Based Measures of Behavior and Psychopathology

To provide a thorough phenotypic assessment of behavior and psychopathology, we gathered information from all SCA and XY control participants using several questionnaire-based, parent-report measures spanning diverse domains of childhood psychopathology, including features that are typically observed in ASD (e.g., Social Responsiveness Scale – Second Edition, SRS-2 [40]), obsessive compulsive disorder (e.g., Obsessive Compulsive Inventory – Revised, OCI-R [41]), motor coordination disorder (e.g., Developmental Coordination Disorder Questionnaire, DCDQ [42]), ADHD (e.g., Conners 3 [43]), impulsivity (e.g., Barratt Impulsiveness Scale, BIS [44]), conduct/dissocial disorders (e.g., Antisocial Process Screening Device, APSD [45]), and aggression (e.g., The Children’s Scale of Hostility and Aggression: Reactive/Proactive, C-SHARP [46]). To supplement these domain-specific instruments, we also included the Child Behavior Checklist (CBCL) [47] and Strengths and Difficulties Questionnaire (SDQ) [48] as multidimensional measures of childhood psychopathology. Thus, our full battery of questionnaire-based measures provided a set of 66 continuous variables (henceforth “scales”) which collectively spanned most major domains of psychopathology in youth. In this report, we use the term “psychopathology” to encompass all of the variables derived from our questionnaire-based behavioral measures (acknowledging that some measures relate to neurodevelopmental features such as motor coordination). The redundancy between scales in the battery was an intentional aspect of our study design as it allows for the same construct to be captured by different instruments and provides a means of assessing whether observed cross– scale correlations are organized according to shared phenomenology as opposed to more superficial features such as instrument of origin. A full list of these scales and their abbreviations is given in **Supplementary Table 1**.

### Cognitive and Adaptive Behavior Assessments

The Wechsler Preschool and Primary Scale of Intelligence, Fourth Edition, Wechsler Intelligence Scale for Children, Fifth Edition, or Wechsler Adult Intelligence Scale, Fourth Edition were used to assess intelligence. If the participant had been tested using the aforementioned Wechsler scales within 1 year (n=1 XXY/KS, n=4 XYY), the Wechsler Abbreviated Scale of Intelligence, Second Edition was used instead. We used these instruments to generate a single Full Scale Intelligence Quotient (henceforth “IQ”) score for each study participant. Adaptive behavior was measured via the Vineland Scales of Adaptive Behavior – Third Edition [49] in the XXY/KS cohort (focusing on the composite score, VABC). The Vineland Scales of Adaptive Behavior – Second Edition was used to assess adaptive behavior in the XYY cohort [50].

### Statistical Analysis

Categorical variables were described using counts and rates, and continuous variables using means and standard deviations. Chi-squared tests and t-tests were used for comparison of categorical (DSM-5 diagnoses) and continuous (questionnaire-based dimensional traits) variables (respectively) as a function of SCA group (XXY/KS vs XYY). An overview of the analytic workflow in this study is provided in **Figure 1** and detailed further below.

**Figure 1.**
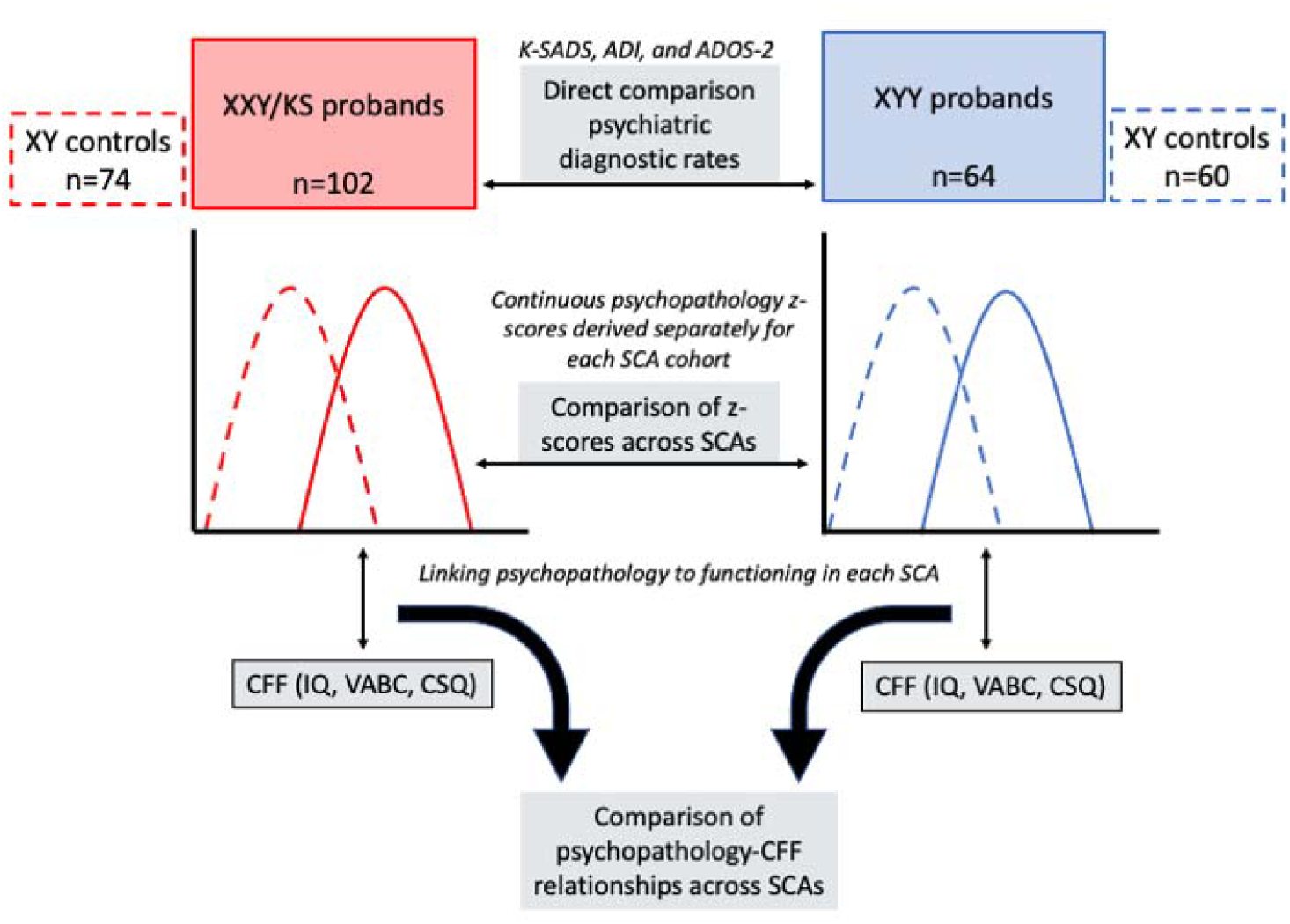
Schematic of analytic workflow. Gold-standard diagnostic assessments were completed for each SCA group and diagnostic rates were compared directly across groups. For each SCA group, z-scores were calculated per psychopathology scale based on the distribution of raw scores in each respective XY control group (middle row). These z-scores were then related to each CFF (IQ, VABC, CSQ) using linear regression (bottom row). The outputs of these regression models were compared using correlations and orthogonal regression to determine if the coupling between psychopathology and CFFs differed as a function of SCA group.

### Diagnostic Rates in XXY/KS and comparison with XYY syndrome

The current report presents DSM-5 diagnostic rates for XXY/KS and a comparison of DSM-5 diagnostic rates between pre– vs. postnatally diagnosed XXY/KS subgroups. Linear regression was used to relate variation in the total number of diagnoses across XXY/KS individuals to three cognitive and functional features (CFFs): IQ, adaptive behavior (VABC), and caregiver strain (CSQ). We compared diagnostic rates in XXY/KS with those previously reported in XYY syndrome [29] using chi-square tests (for all instances with ≥5 expected observations per cell [51]). We used linear regression to test if karyotype significantly modifies relationships between diagnostic count and CFFs (diagnostic count as a dependent variable modeled by the main and interactive effects of karyotype and CFF).

### Questionnaire-Based Measures of Behavior and Psychopathology in XXY/KS

We profiled the effect of XXY/KS on 66 dimensional measures of psychopathology as previously described for XYY [29]. Since not all of the originating questionnaires provided published normative sample scores, we used the following procedure to bring all XXY/KS scores into a common reference frame relative to score distributions in the 74 age-matched XY controls. We first tested for the presence of a statistically significant difference in age effects on scale scores between XXY/KS case and XY controls. Where such age-by-group interactions were found, we re-expressed observed scores for all XXY/KS and XY individuals as standardized residuals from predicted scores for their age given by a general linear model for score as a function of age estimated in our XY control cohort. In the absence of such age-by-group interactions, we expressed XXY/KS scores for a scale as z-scores using the distribution in XY controls. This procedure expressed the score for each scale in each XXY/KS individual as a z-score relative to the score distribution observed in XY controls.

The magnitude and statistical significance of z-score differences between XXY/KS and XY groups were identified using linear models with scale score as a continuous dependent variable and SCA group as a categorical predictor – for the full XXY/KS group as well as the subgroup of prenatally diagnosed individuals. For each scale, we also computed standardized regression coefficients for the relationship between score variation in the XXY/KS group and CFFs, and timing of XXY/KS diagnosis (prenatal vs. postnatal). All statistical tests for dimensional measures in XXY/KS were Bonferroni-adjusted for multiple comparisons across scales (adjusted p– value: 0.05/66=0.00076).

### Comparing Dimensional Measures of Psychopathology Between SCAs

The data collection and preprocessing steps described above were identical for both the current newly-described XXY/KS cohort and the previously described XYY cohort [29] yielding individual-level z-scores for 66 dimensional measures in 102 XXY/KS and 64 XYY participants. We first compared SCA groups by binarizing each of these score distributions using a classical z-score cutoff of >2 (to identify SCA individuals with elevated levels of psychopathology above the 97.5^th^ centile of their respective XY distribution), and using chi-squared tests (or Fisher tests when expected observations per comparison <5 [51]) to compare the proportion of individuals above this cutoff in XXY/KS vs. XYY groups (correcting for multiple comparisons across scales with an adjusted p-value of 0.05/66=0.00076). We next compared the full distribution of z– scores between XXY/KS and XYY groups for each scale using linear models with z– score as the dependent variable and group (XXY/KS vs. XYY) as the independent variable. This procedure estimated the magnitude (beta coefficient for the group term, henceforth “Δz-score”) and statistical significance (adjusted p-value: 0.05/66=0.00076) of karyotype group differences in mean score for each measured dimension.

### Comparing Profiles of Psychopathology and Coupling with CFFs Between SCAs

We used correlation and regression analyses to directly compare the profile of mean z– scores across all 66 measures between XXY/KS and XYY groups. The Pearson correlation coefficient for these 66 measures between groups estimates the strength of the linear relationship between psychopathology profiles in the SCA groups. We also examined the relationship between mean z-score profiles in XXY/KS and XYY using an orthogonal regression framework to where the variation in mean z-scores across scales in one SCA is modeled as a function of that in the other SCA. This approach: provides a formal model enabling prediction of the mean effect of one SCA on a dimension of psychopathology from the mean effect in another SCA; tests for deviation of this relationship from the null hypothesis of identical score profiles (i.e. a fit line with an intercept of 0 and a slope of 1); and, quantifies any deviations of individual subscales from the general relationship (i.e. orthogonal residuals of individual subscales from the regression fit line). We used orthogonal, rather than ordinary least squares regression for these purposes because the former provides stable model coefficients regardless of arbitrary decisions regarding which karyotype group scores are treated as the dependent vs. the independent variable [52].

## RESULTS

### Participant Characteristics

Participant characteristics for the newly reported XXY/KS cohort (102 XXY/KS and 74 age-matched XY controls) are detailed in **Table 1**, which also reproduces demographic information for the previously reported XYY cohort (64 XYY individuals and 60 age– matched XY controls, [29]). XXY/KS cases and XY controls showed significant differences in IQ and socioeconomic status, but not in age. There were no statistically significant differences between pre– and postnatally diagnosed XXY/KS subgroups (n=38 and n=64, respectively) in age, IQ, VABC, or CSQ scores. Compared to their XYY counterparts, the XXY/KS group had a higher mean IQs (t=3.15, p=.002) and their caregivers reported lower mean levels of caregiver strain on the CSQ (t=-.2.57, p=.012). The two SCA groups did not differ in mean VABC scores (t=.52, p=.6).

**Table 1.**
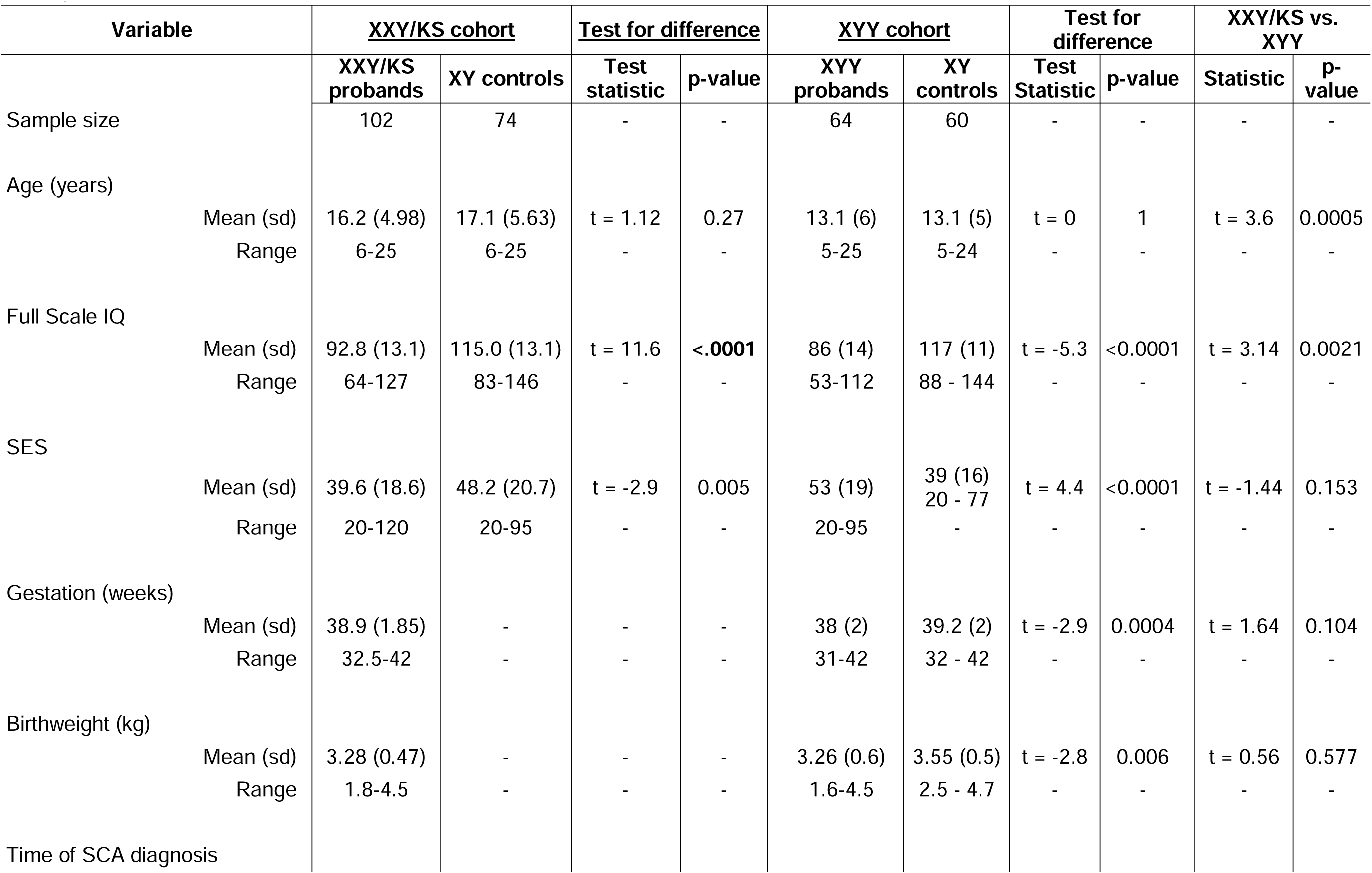
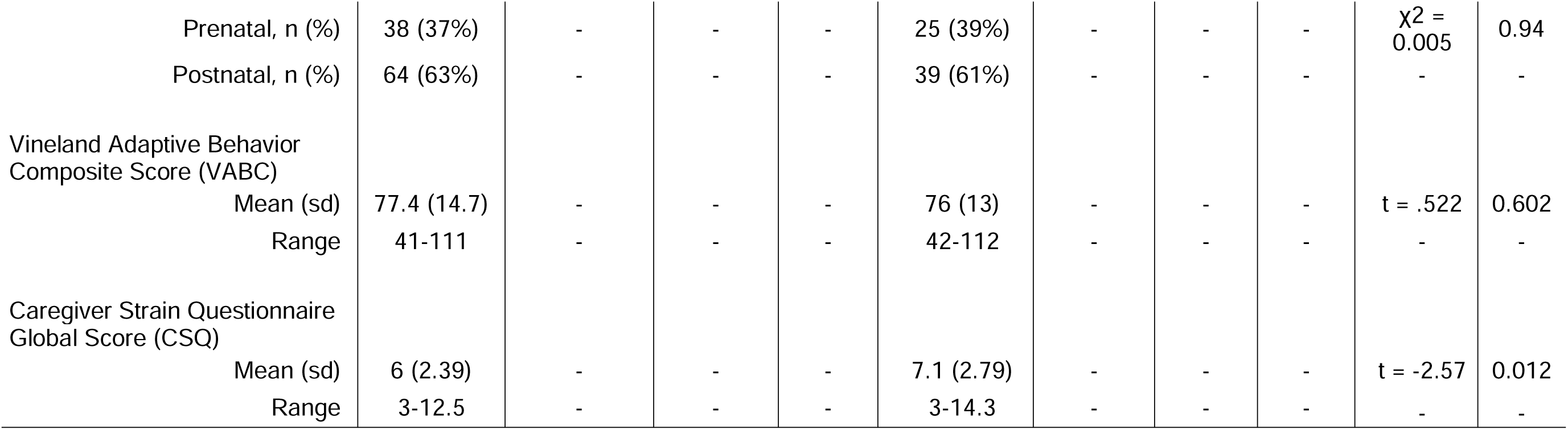
*Participant characteristics*

### Psychiatric Diagnoses in XXY/KS and Comparisons with XYY

As revealed by the K-SADS, ADI-R, and ADOS-2, the XXY/KS cohort displayed high rates of psychiatric morbidity, with 87% of this group ever having had a DSM-5 diagnosis at the time of assessment. Neurodevelopmental disorders (NDDs) was the most prominent diagnostic category (79% ever having a lifetime diagnosis), followed by anxiety disorders (33%), elimination disorders (27%), and mood disorders (18%). The three most prevalent individual diagnoses within XXY/KS were ADHD (65%, of which 82% met inattentive subtype criteria), ASD (34%), and enuresis (26%) (see **Table 2** for complete diagnostic summaries). The postnatally diagnosed subgroup did not show statistically significant enrichment for any diagnostic category. Regression analyses revealed that the number of lifetime psychiatric diagnoses in XXY/KS was associated with significantly greater CSQ scores (β=.67, p=.0002) and significantly lower VABC scores (β=-5.36, p<.00001), but was not significantly associated with IQ (β=-1.56, p=.10).

**Table 2.**
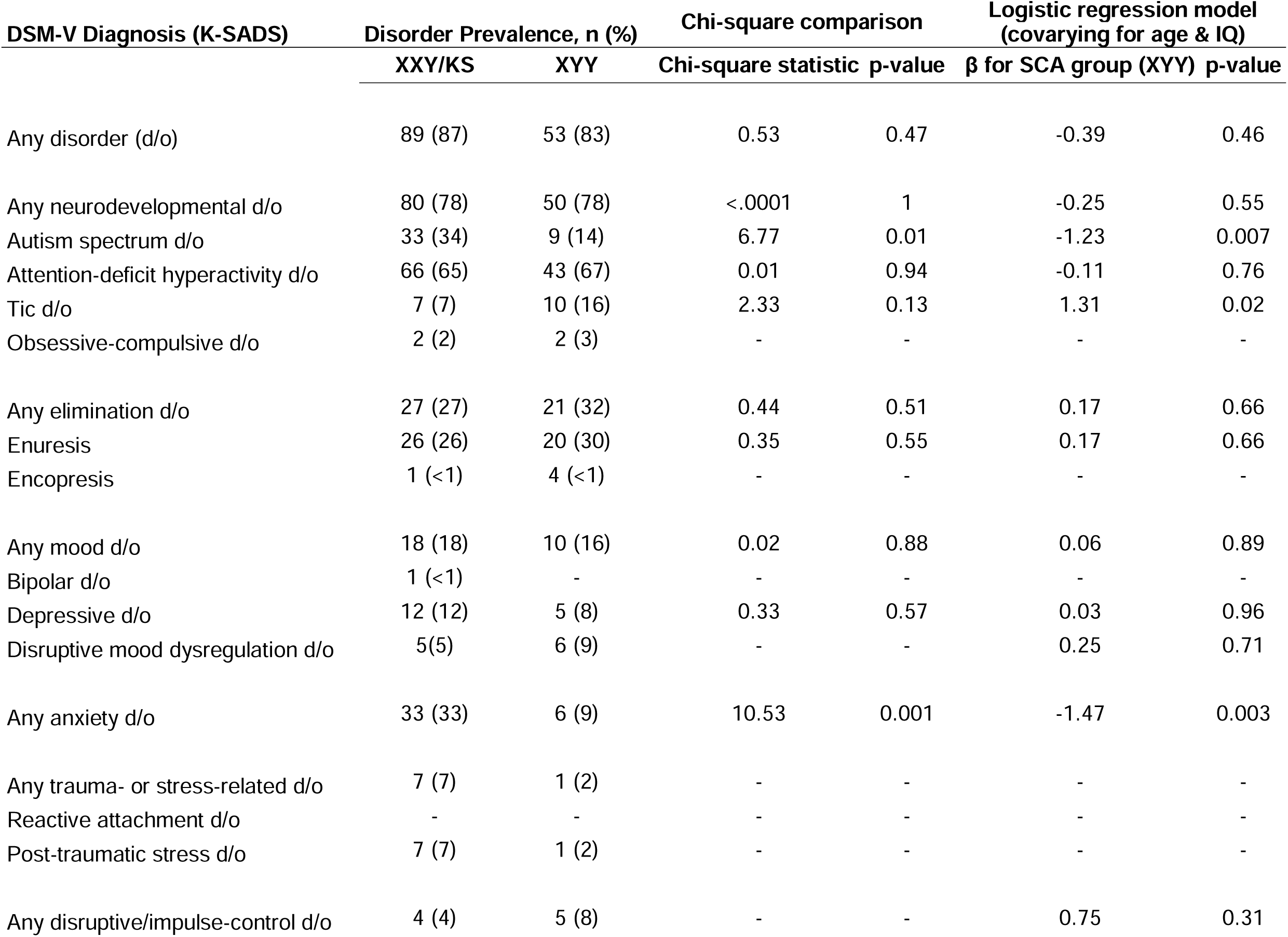
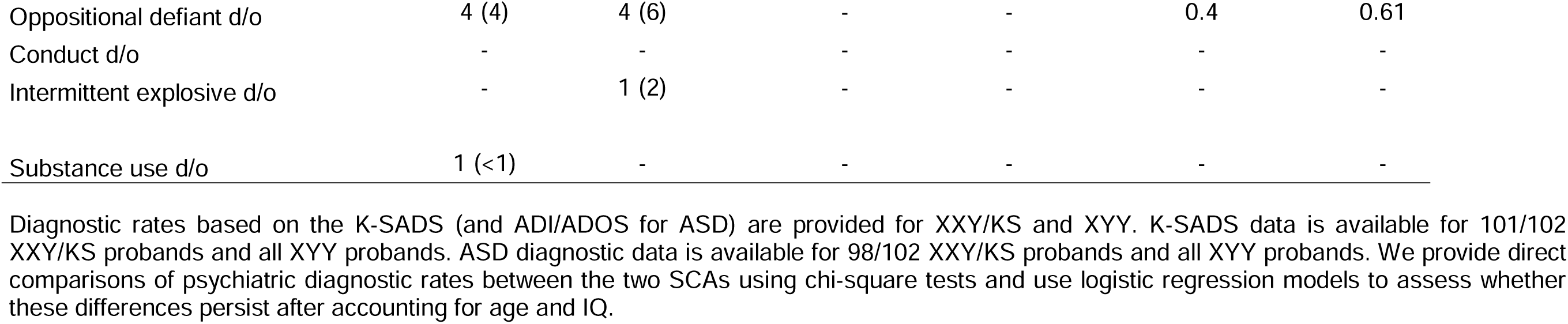
Psychiatric diagnostic rates in XXY/KS and XYY

Comparison of diagnostic rates between XXY/KS and XYY identified significant differences for anxiety disorders, which were overrepresented in XXY/KS (X^2^=10.53, p=.001). ASD was nominally more prevalent in XXY/KS, but did not survive correction for multiple tests (X^2^=6.77, p=.01). Both SCA groups demonstrated high rates of comorbidity, with the modal number of diagnoses being two, and over half of each SCA group having 2 or more diagnoses by the time of assessment. Interaction analyses revealed that the relationships between number of lifetime diagnoses and our three CFFs did not significantly differ between the two karyotype groups. However, the linear relationship between number of diagnoses and IQ was significant only in XY

### Dimensional Scores of Psychopathology in XXY/KS

The 12 instruments used in this analysis yielded a total of 66 overlapping scales spanning several dimensions of childhood psychopathology. The number of XXY/KS participants with available scores for a given scale ranged between 38 and 101 due to differences in the applicable age-range for each questionnaire, but the XXY/KS and XY groups with available scores for each scale did not differ in mean age (**Sup Table**).

For each measured scale, we expressed individual-level scores in the XXY/KS group as z-scores relative to the respective score distribution observed in the XY control group. Doing so allowed us to rank all 66 measured dimensions by their relative disruption in the XXY/KS group (median Δz-score, difference between XXY/KS and XY) in a single combined box plot visualization (arranged by median z-score, **Figure 2A**). The resulting ranking of scales was generally organized by psychopathological domain rather than instrument, with greatest score elevations observed in measures of ASD– related traits (e.g., SRS-2 social cognition subscale, Δz-score=2.8), social deficits (e.g., SRS-2 social communication subscale, Δz-score=2.4), and mood/anxiety symptoms (e.g. CBCL internalizing symptoms subscale Δz-score=3.15). Intermediate rankings were generally observed for attention problems (e.g., Conners 3 inattention subscale, Δz-score=1.9), motor coordination deficits (e.g., DCDQ total score, Δz-score=1.6), executive dysfunction (e.g., Conners 3 executive functioning subscale, Δz-score=1.71), and disruptive behaviors (e.g., CBCL rule-breaking behavior subscale, Δz-score=1.78). Minor, and often statistically insignificant, elevations were seen in scales assessing obsessive-compulsive features (e.g., OCI ordering subscale), aggressive behaviors (e.g., C-SHARP physical aggression subscale), and antisocial tendencies (e.g., APSD callous-unemotional subscale). More than two thirds of all measured scales (46/66, 70%) showed statistically significant score elevations in XXY/KS compared to XY controls after Bonferroni correction. Within XXY/KS, scores on most scales tended to be higher in postnatally vs. prenatally diagnosed subgroups, although no individual scale showed significant score differences between these subgroups after Bonferroni correction for multiple comparison across scales (**Fig 2D**). A majority of these elevated scales (28/46, 61%) remained significantly elevated in XXY/KS as compared to XY controls when restricting analyses to prenatally diagnosed XXY/KS individuals (**Fig 2E**), with similar magnitudes observed across scales. It should be noted that scale elevation was not entirely attributable to differences in IQ, as 19 scales remained significantly elevated after adjusting for variation in IQ in a separate model (**Supplementary Table 1)**.

**Figure 2.**
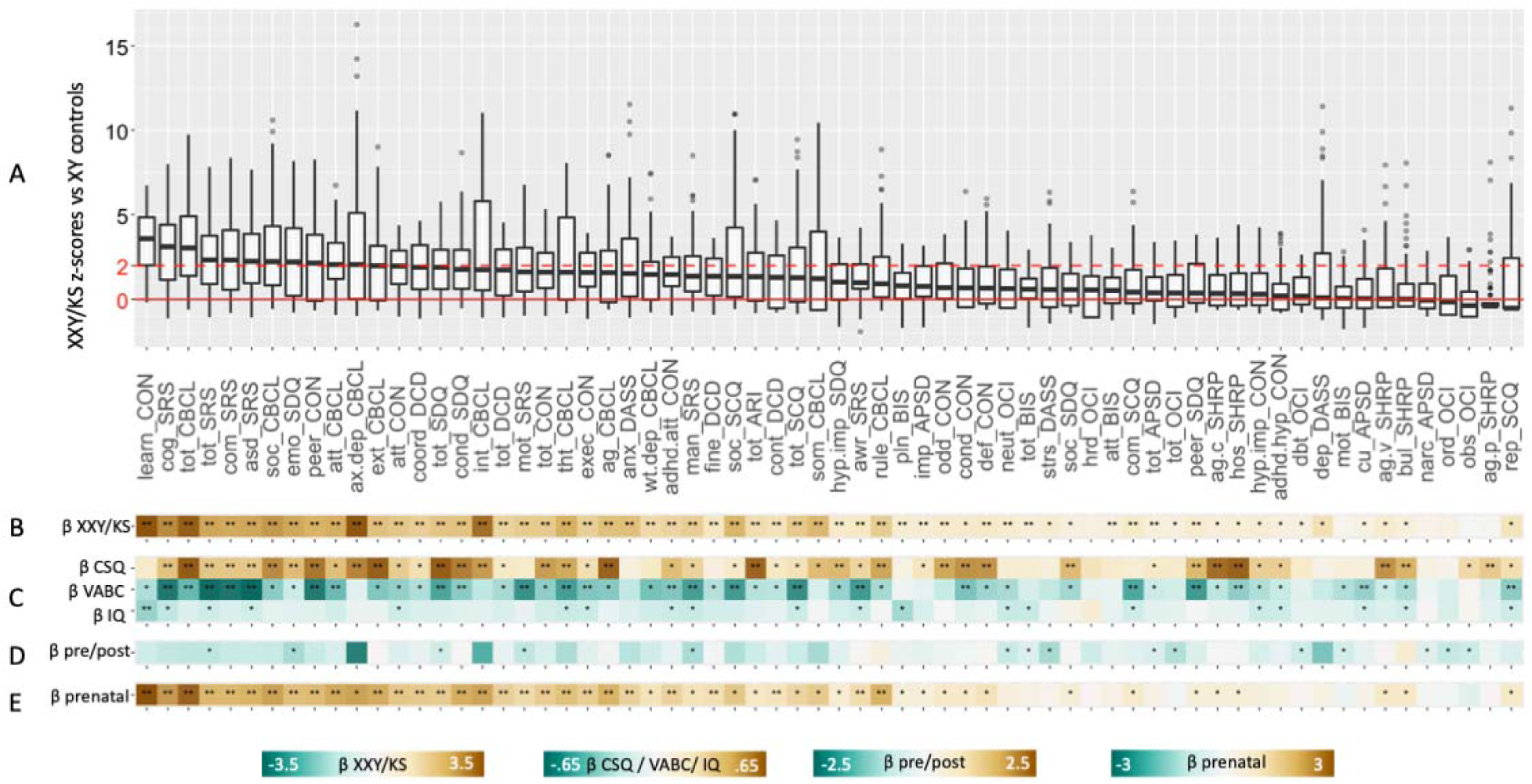
Profiling symptoms in XXY/KS across 66 scales. (A) Boxplots showing distribution of z-scores in XXY/KS for 66 different subscales derived from 12 questionnaires. Horizontal red line shows mean score in XY controls, and dashed horizontal line shows a 2 standard deviation elevation in scores relative to the observed distribution in XY males. **(B)** Standardized beta coefficients for the effects of XXY/KS (vs. XY) on each scale. **(C)** Standardized beta coefficients for regression models predicting variation in IQ (Full-Scale IQ), VABC (Vineland Adaptive Behavior Composite Scale), and CSQ (Caregiver Strain Questionnaire) scores as a function of variation in each measure of psychopathology. **(D)** Standardized beta coefficients for association between time of XXY/KS diagnoses (postnatal vs. prenatal) and scores on each measure of psychopathology. **(E)** Standardized beta coefficients for the effects of XXY/KS (vs. XY) on each scale when analysis is restricted to the subset of XXY/KS individuals who were diagnosed prenatally. For **B**–**E**, a single asterisk indicates statistical significance at uncorrected p=0.05, and a double asterisk at Bonferroni– adjusted p<.00076.

We next used linear regression to relate interindividual variation of scores within each measured dimension of psychopathology in XXY/KS to accompanying variation in each of the three CFFs of interest (**Figure 2B**). After correction for multiple comparisons across scales, learning problems on the Conners 3 was the only scale significantly correlated with IQ in XXY/KS (β=-.18, p=.0004), whereas variation in VABC and CSQ were significantly associated with scores on numerous dimensions of psychopathology (23 and 29 scales respectively). ASD-related, social, attentive, and externalizing features tended to have the most robust relationships with VABC and CSQ. Interestingly, the scales most strongly associated with VABC and CSQ were not necessarily among the most impacted in XXY/KS. The hostility subscale of the C– SHARP, for instance, showed robust associations with VABC and CSQ scores (β=.33 and .61, p<.0005 and .0001, respectively), despite not being significantly elevated in the overall XXY/KS cohort.

### Comparing Scores on Individual Dimensions of Psychopathology Between XXY/KS and XYY Syndrome

We took two complementary approaches to comparing scores on individual dimensions of psychopathology between XXY/KS and XYY. First, given the wide distribution of scores on psychopathology dimensions in XXY/KS (**Figure 2B**) and XYY (Raznahan et al., 2023), we used a z-score cutoff of 2 to define the proportion of individuals in each group with severely elevated scores beyond the 97.5^th^ percentile of the XY score distribution for each scale (**Figure 3A**). This proportion was larger in XYY than XXY/KS for most scales (42/66, 64%) at a nominal significance level, and these differences were statistically significant after Bonferroni correction for 18% (12/66) of scales. The largest of these disparities in the rate of extreme scores between XYY and XXY/KS were seen for SRS-2 restricted interests and repetitive behaviors (mannerisms), SDQ peer problems, and OCI mental neutralizing scales. In contrast, just one scale showed a nominally significant (p<0.05 uncorrected) greater proportion of individuals with severely elevated scores in XXY/KS as compared to XYY: emotional problems on the SDQ.

**Figure 3.**
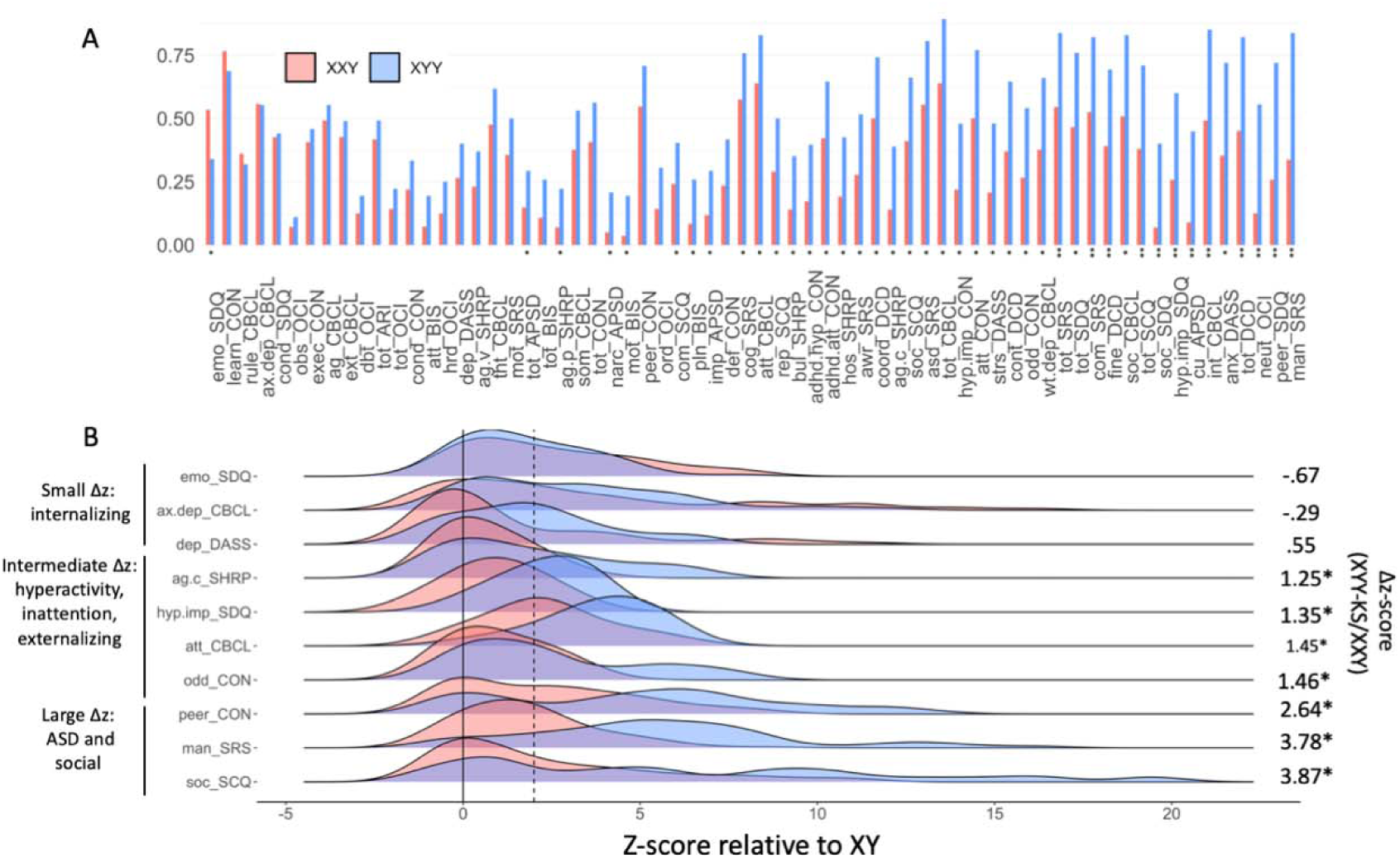
**Direct comparisons of extreme score rates for 66 dimensional measures of psychopathology between XXY/KS and XXY syndrome, with illustrative score distributions for selected traits**. **(A)** Bar graph displaying the proportion of individuals with a z-score > 2 SDs above their respective control group (XXY/KS = red, XYY = blue). Subscales are arranged in order of ascending proportion difference (XYY– XXY/KS). One asterisk (*) indicates the p-value for the chi-square statistic is less than .05, two asterisks (**) denotes the p-value is less than .00076. **(B)** A selection of density plots illustrating the distribution of z-scores for a selected sample of subscales (XXY/KS = red, XYY = blue). Solid line is at z-score of 0 (indicating average of XY controls), and dashed line is at 2 (indicating 2 standard deviations above XY controls).

Second, we complemented the cutoff based approach above with a comparison of the continuous distribution of z-scores between groups. This approach revealed that 82% (54/66) of scales had a greater mean z-score in XYY than XXY/KS (p<.05), and these differences were significant after Bonferroni correction for 53% (35/66) scales (**Supplementary Table 1**). **Figure 3B** illustrates the distribution of z-scores in XXY/KS and XYY for a selection of scales representing different levels of score disparity. The largest disparities were observed in scales measuring social deficits (e.g., XYY vs. XXY/KS Δz for SRS-2 social communication=2.03, p< .0001) and ASD-related traits (e.g., XYY vs. XXY/KS Δz for SRS-2 restricted interests & repetitive behavior (mannerisms) subscale=3.78, p<.0001), with moderate differences seen in measures of inattention, hyperactivity, and externalizing behaviors (e.g., XYY vs. XXY/KS Δz for SDQ hyperactivity/impulsivity subscale=1.35, p<.0001). Conversely, the differences in effect sizes for some mood– and anxiety-related scales between the SCAs were negatively signed, but relatively small in magnitude and often failed to reach statistical significance (e.g., XYY vs. XXY/KS Δz for SDQ emotional problems=-0.67, p=.08). Given that the XXY/KS and XYY cohorts differed in mean age and IQ, we also assessed psychopathology differences between the SCAs in a separate model covarying for these variables (results shown in **Supplementary Table 1**). While some of the originally observed differences no longer reached the significance threshold in this adjusted model, a majority (24/35, 69%) remained significant with only slight attenuation in their effect sizes. Taken together, these comparisons suggest that most domains of psychopathology are more severely impacted in XYY than XXY/KS (the disparity being most marked for social and attentional problems), with the notable exception of internalizing symptomatology, which stands out for being comparably impacted in XXY/KS and XYY.

### Comparing Profiles of Psychopathology and the Coupling of Psychopathology with CFFs Between XXY/KS and XYY Syndrome

We next sought to combine information across all measured scales to directly compare the relative ranking of scales by effect-size (i.e., penetrance – quantified as mean z– score) between XXY/KS and XYY. In contrast to the comparisons of individual scales above (**Figure 3**), these analyses assess the concordance in psychopathological *profile* (i.e., vector of mean z-scores across scales) between the SCAs. We quantified this concordance using two complementary approaches: Pearson correlation and orthogonal regression.

The cross-scale correlation in effect sizes between XXY/KS and XYY was strong (r=0.75, p<.00001, see **Figure 4A**). Thus, we observe a strong positive correlation in the impact of XXY/KS and XYY across different domains of psychopathology – with general concordance between karyotypes in the domains most vs. least impacted. However, the same correlation coefficient could arise through very different scenarios which can be disambiguated by regression analyses, such as there being a consistent offset by which all scores are shifted in one SCA relative to the other, or there being a variable offset across scales.

**Figure 4.**
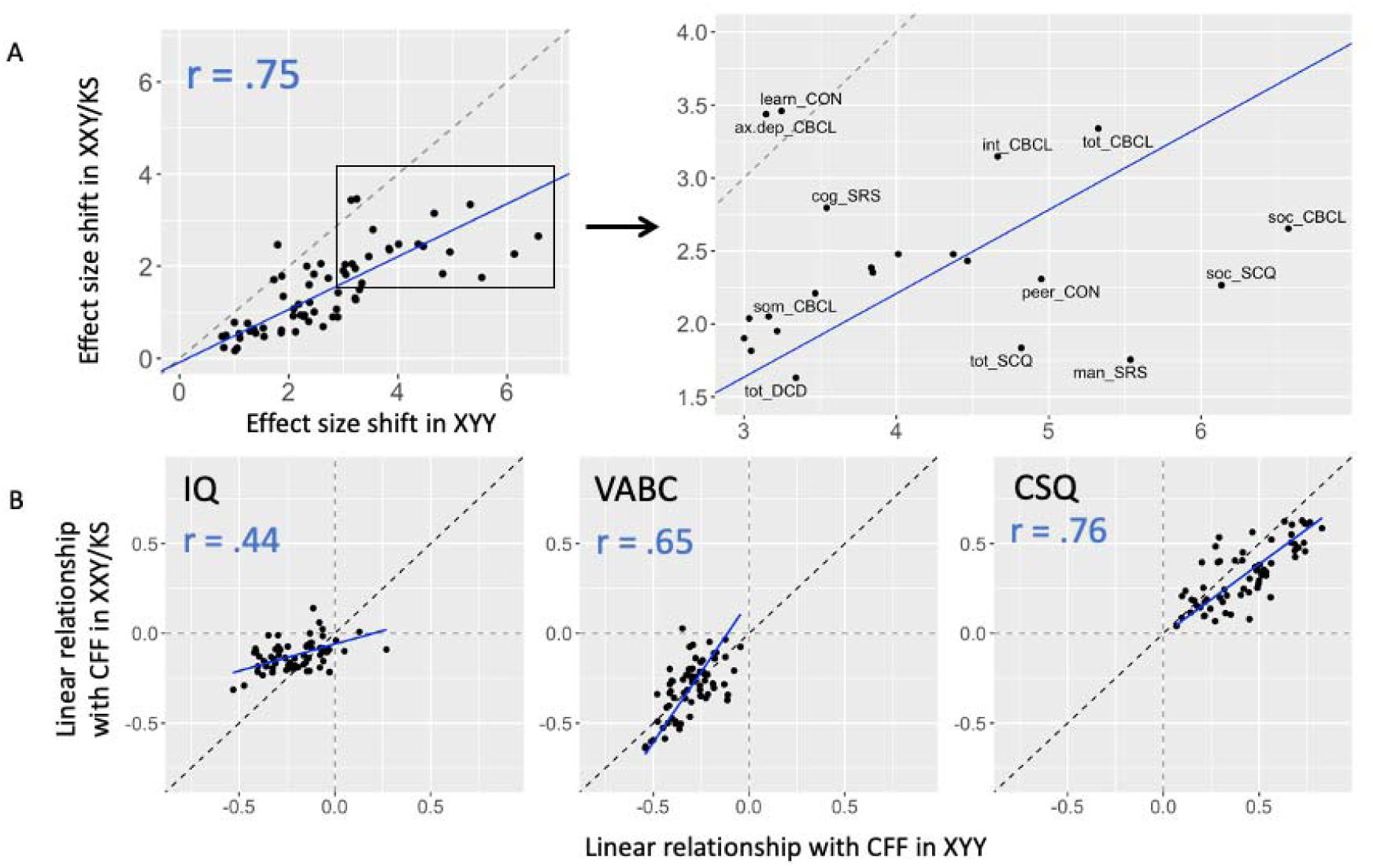
Orthogonal regression modeling to assess the concordance of psychopathological profiles and CFF coupling across SCAs. (**A**) Scatterplot and orthogonal regression modeling the relationship between mean scores in XXY/KS and XYY for each scale. A point’s position along the x– and y-axes represent its mean effect in XYY and XXY/KS, respectively. A subset of the graph (boxed) is expanded to illustrate the position of scales which deviate the most drastically from the regression slope. (**B**) Scatterplot and orthogonal regression modeling the relationship between scales’ relationship with CFFs (IQ, VABC, and CSQ) in XXY/KS and XYY. A point’s position along the x– and y-axes represent the scale’s linear relationship with the CFF in XYY and XXY/KS, respectively.

Orthogonal regression modeling of the relationship between mean scale scores in XXY/KS and XYY revealed a systematic shift of scale scores in XXY/KS relative to XYY, such that the regression fit line deviated from the identity line which would define perfect congruence in scale scores between the two SCAs. Specifically, the slope of the fit line modeling XXY/KS scores as a function of those in XYY was significantly less than the identity line slope of 1 (slope=.57, p <.00001 vs. null hypothesis of slope=1) – indicating there is a systematic tendency for scores to be less severe in XXY/KS than XYY, and that this disparity becomes greater for scales that are more severely impacted in XYY. In other words, to the extent that a given aspect of psychopathology is impacted by XXY/KS – it tends to be more impacted by XYY. Estimation of this slope also makes it possible to quantify the deviation of individual scales from model predictions. Of note, we observed the largest negative residuals – that is, scales with most marked increase in severity within XYY relative to XXY/KS – for the mannerism subscale of the SRS (orthogonal residual=-1.33), the communication subscale of the SCQ (orthogonal residual=-1.17), and the social problems subscale of the CBCL (orthogonal residual=– 1.03). Conversely we observed the largest positive residuals for the anxious-depressed subscale of the CBCL (orthogonal residual=1.72), the learning subscale of the Conners (orthogonal residual=1.68), and the emotional problems subscale of the SDQ (orthogonal residual=1.52) – indicating these scales break the overall trend of impairments tending to be more severe in XYY than XXY/KS. Indeed, these scales fell close to the identity line indicating near-identical severity between the two SCAs. Thus, presence of an extra X– and Y-chromosome seems to have a surprisingly similar capacity to impact mood features given that most other domains are more sensitive to a supernumerary Y– than a supernumerary X-chromosome.

Finally, we applied the same pairing of correlation and regression analysis to compare XXY/KS and XYY for the coupling between psychopathology and CFFs. These analyses asked if the two SCAs were similar in the relationships they each showed between psychopathology and CFFs. Correlation analysis revealed that the between– SCA congruence in psychopathology-CFF coupling was greatest for CSQ (r=.76, p<.00001), intermediate for VABC (r=.65, p<.00001), and weakest for IQ (r=.44, p=.0002). Thus, relationships between psychopathology and caregiver strain are highly conserved between XXY/KS and XYY, whereas we observe a weaker conservation of the relationships between psychopathology and IQ (**Figure 4B**). Applying orthogonal regression to the coupling of psychopathology and CFFs provides further insight into the connectivity of different behavioral features across the SCAs. For example, although no significant differences were observed for any individual scale in its relation with IQ between SCAs, the output of the regression reveals a depressed slope (slope=.3, p <.00001 vs. null hypothesis of slope=1), indicating that negative relationships between psychopathology and IQ tend to be stronger in XYY than in XXY/KS. The same analytic approach also suggests a stronger overall psychopathology-CSQ coupling in XYY as compared to XXY/KS (slope=0.78, p=.0005 vs. null hypothesis of slope=1).

## DISCUSSION

As detailed below, by applying several complementary analytic approaches to deep phenotypic data, our study design substantially advances understanding of X– and Y-chromosome dosage effects on psychopathology and extends the toolkit for securing similar advances in other GDDs.

### Elevated rates of psychiatric diagnoses in XXY/KS

By systematically profiling psychiatric diagnoses using gold-standard diagnostic instruments in enrolled individuals, we find elevated rates for most psychiatric diagnoses in XXY/KS, with highest rates for ASD, ADHD, and anxiety disorders. High rates of psychiatric comorbidity were observed in XXY/KS, and the degree of this comorbidity was shown to relate to caregiver strain and deficits in adaptive behavior, meaning that diagnostic burden may provide insight into an individual’s levels of overall functioning, extending beyond psychiatric symptoms. Similarly broad increases in diagnostic rates have been reported by prior studies in both self-selected and population-based XXY/KS cohorts [21, 53]. Of note, the diagnostic rates observed in self-selected XXY/KS cohorts by the present study and Bruining et al. (2009) imply hazard ratios for increased psychiatric risk that are higher than those recently published in the only available population based XXY/KS cohort [21]. The same statement holds for self-selected vs. population based XYY cohorts. These inflations of estimated penetrance likely reflect ascertainment bias in clinical vs. population-based GDD cohorts, as has been previously shown for other more extensively studied GDDs such as 22q11.2 deletion [54]. However, our observation of substantial diagnostic comorbidity and subthreshold symptomatology in both XXY/KS (current report) and XYY syndrome [29] stresses the importance of examining fine– grained dimensional measures of psychopathology in GDDs, and such detailed measures are typically not available in population-based datasets. We therefore paid particular attention to such dimensional measures in our study while using comparisons between pre– and postnatally ascertained XXY/KS individuals to probe for potential ascertainment bias effects (see below).

### Dimensional profiling of psychopathology in XXY/KS

By analyzing scores across 66 dimensional measures, we provide a systematic ranking of psychopathology domains by the severity of their score elevations in XXY/KS. We find that carriage of a supernumerary X-chromosome in males appears to target certain domains of behavior (e.g.m social functioning and mood) while leaving others relatively unaffected (e.g., antisocial and obsessive-compulsive features). Crucially, this ranking of behavioral domains by their vulnerability to XXY/KS is highly conserved between subsets of our cohort with different likelihoods of ascertainment bias (prenatally vs. postnatally diagnosed individuals), suggesting that it is an accurate and generalizable signature of strengths and weaknesses in XXY/KS. Importantly, we show that top-ranking scales are not necessarily those that are most tightly coupled with interindividual variation in cognitive ability, adaptive behavior and caregiver strain within XXY/KS. Taken together, these behavioral insights from fine-grained dimensional data can inform future clinical and neurobiological research in XXY/KS. In a clinical context, insight into the behavioral signatures of supernumerary X-chromosome carriage may help to expedite psychiatric or neuropsychological assessment. For example, given that mood and social features pose a particular vulnerability in XXY/KS, these domains should be prioritized by clinicians during psychiatric or developmental assessment. Relatedly, awareness of the domains of psychopathology that are most tightly coupled to adaptive behavior (e.g., inattention) or caregiver strain (e.g., hostility) may provide clinicians the opportunity to pinpoint behavioral tendencies which associate in a specific manner with these functionally relevant features. Although these data are cross-sectional in nature, and therefore cannot address causality, they offer promising first steps in identifying intervention targets whose treatment may subsequently improve adaptive skills or alleviate caregiver stress.

### Deep Phenotypic Behavioral Comparison Between XXY/KS with XYY

For both categorical diagnostic outcomes and dimensional measures, we observe a shared capacity of XXY/KS and XYY to induce a broad increase in risk for psychopathology. Direct comparison of diagnostic rates highlights a significantly elevated risk of anxiety disorders in XXY/KS relative to XYY, and we find that extreme scores on dimensional measures of depression and anxiety are also more prevalent in XXY/KS than XXY (albeit not statistically significantly so). Several depression and anxiety scales stand out in cross-scale correlation of mean scores between XXY/KS and XYY for not cohering with the general pattern of greater symptom severity in XYY than XXY/KS (i.e., comparable elevations across the two SCAs). Taken together these findings suggest that carriage of an extra X-chromosome in males imparts a greater elevation in relative risk for internalizing symptoms as compared to carriage of an extra Y-chromosome. Conversely, most other dimensions of psychopathology tend to be more severely impacted in XYY vs. XXY/KS. This disparity is particularly pronounced for measures of social impairment, inattention, hyperactivity, and externalizing behaviors. Knowing that some domains of psychopathology can be differentially impacted by XXY/KS vs. XYY may help clinicians tailor their clinical assessments according to SCA karyotype, and also motivates research into the biological bases for these differential effects of the X– and Y-chromosome on human behavior (see below). More generally, the analytic approach we apply here to two SCAs can be generalized to inform tailored clinical assessment and neurobiological research in other GDDs.

### X-and Y-Dosage Effects on the Coupling of Psychopathology and Cognitive & Functional Features (CFFs)

For XXY/KS and XYY – as for most GDDs – increased risk for psychopathology is accompanied by co-occurring impairments in cognition [1, 55] and adaptive functioning [56, 57] along with potential increases in caregiver strain [58]. Because impairments in such CFFs are well known to correlate with levels of psychopathology amongst genetically unselected youth [59–61], we tested for the presence and genotype-dependence of such correlations in XXY/KS and XYY. Our results suggest that some links between psychopathology and CFFs are highly stable between different GDDs whereas others appear to be more GDD-specific. For example, adaptive behavior and caregiver strain are tightly coupled to multiple domains of psychopathology, with the strength of these relationships being highly correlated (r>0.6) between XXY/KS and XYY. This coupling across SCAs could partially reflect measurement issues if similar questions appear on ratings scales measuring psychopathology and adaptive behavior (e.g. items probing difficulty in peer relationships). Alternatively, this concordance in coupling may also reflect common pathways to CFFs through psychopathology (e.g. externalizing behaviors driving caregiver strain, regardless of the behavior’s genetic etiology) [62]. These results may help clinicians target their assessment to particular domains of psychopathology when seeking to contextualize adaptive functioning and caregiver strain in both XXY/KS and XYY (and potentially other GDDs once the necessary comparative data on psychopathology-CFF coupling become available). In contrast to these findings for adaptive functioning and caregiver strain, the mapping between psychopathology and IQ is only moderately correlated between XXY/KS and XYY. Most dimensions of psychopathology are more strongly coupled to IQ variation in XYY than XXY/KS, and this is particularly true for measures of ASD traits and impulsivity. Our cross-sectional and observational study design cannot discern the causal basis for these findings but helps to refine several testable hypotheses. For example: greater cognitive impairments may partly drive greater psychopathology [63, 64] and this effect may be more pronounced in XYY than XXY/KS; greater psychopathology may drive lower performance in IQ tests and this effect may be stronger in XYY than XXY/KS; there may be nonlinear associations between psychopathology and cognitive impairment so that correlations are stronger at the higher levels of psychopathology observed in XYY as compared to XXY/KS (however, see Mahony et al., 2023); or, the neurobiological pathways underpinning psychopathology and cognitive impairment show greater overlap in XYY than XXY/KS.

### Mechanisms for Observed Convergences and Divergences Between XXY/KS and XYY

The comparative findings above help to differentiate those clinical aspects that are shared between XXY/KS and XYY (e.g., the ranking of behavioral domains by impairment or the coupling between psychopathology and caregiver strain) from those that appear to be more karyotype-specific (e.g. symptoms in affective, social and attentional domains, or the coupling between psychopathology and IQ). The potential mechanistic bases for these convergences and divergences can be considered across several levels of biological analysis spanning from the proximal changes in gene expression caused by alterations in chromosome dosage through to downstream changes in brain structure and function.

At the genomic level of analysis, phenotypic similarities between XXY/KS and XYY may reflect the effects of dosage alteration for those genes that are shared between the X– and Y-chromosome and show similar expression changes in both aneuploidies: pseudoautosomal region (PAR) genes and X-Y gametologs [32, 33]. We speculate that X-Y gametologs may be particularly important for the shared effects of X– and Y-chromosome aneuploidy because these genes are widely expressed across the body [65], under strong evolutionary constraints [66], and highly dosage sensitive in SCA across different tissues [32, 33, 67].

Genetic sources of divergent XXY/KS and XYY effects include: altered expression of brain-expressed gene sets that are not shared between the X– and Y– chromosome such as non gametolog X-linked genes that escape X-inactivation; dosage effects of non-genic regulatory motifs that are unique to the X– and Y-chromosome; and changes in the nuclear environment from the gain in heterochromatin that uniquely accompanies inactivation of the second X-chromosome in XXY/KS (not operative in XYY) [68]. Gains in X– vs. Y-chromosome dosages are also known to differentially impact testicular function in males– for example, progressive hypogonadism is seen in XXY/KS but not XYY [69]. It is therefore theoretically possible that some of the distinguishing behavioral features in XXY/KS as compared to XYY (e.g. lower levels of overall psychopathology, relatively greater risk for mood and anxiety issues, weaker coupling between psychopathology and IQ) could reflect secondary effects of testicular dysfunction on the brain [70, 71]. However, links between hypogonadism and mental health are understudied in XXY/KS, and this is a high-priority area for future research [72].

In contrast to the highly divergent effects of XXY/KS vs. XYY on the testes, these two SCAs appear to have spatially convergent effects on regional brain organization, which may provide a neurobiological basis for shared effects on behavior. For example, after controlling for the divergent effects of XXY/KS and XYY on overall brain size (significantly decreased in XXY/KS and slightly increased in XYY), these two conditions both induce relative reductions in the size of brain regions including right anterior cingulate and right posterior insula, alongside relative increases in the size of regions such as the right orbitofrontal cortex and left medial prefrontal cortex [31]. This distributed set of brain regions displaying similar effects of X– and Y-chromosome gain is known to be particularly important for communication, memory, reward processing, and emotion regulation, which may underpin shared effects of both XXY/KS and XYY on psychopathology [31].

### Caveats and Limitations

Our findings must be considered in light of several caveats and limitations. First, ascertainment biases in clinical GDD cohorts such as those presented here are likely to inflate estimates of penetrance [54]. For diagnostic outcomes, it is possible to minimize ascertainment bias using population-based research in countries with national registries linked to genotyped birth cohorts [21]. But, to date, there are no population-based datasets that offer dense dimensional measures of psychopathology and functioning in GDDs while also fully protecting from ascertainment bias. Although our analysis of such dimensional features is also prone to potential ascertainment bias effects, we systematically qualify our findings with tests for ascertainment bias based on comparison of pre– and postnatally diagnosed XXY/KS (current paper) and XYY [29] individuals. Second, while the combination of diagnostic and dimensional outcomes in our study design provides complementary information, it also illustrates the possibility for apparent inconsistencies between these measurement methods. In the current study, we find ASD rates to be substantially higher (approximately 3-fold) in XXY/KS than in XYY, despite prior comparative literature presenting the opposite pattern [28, 73, 74]. Similarly, we find this difference in spite of our continuous analyses, which suggest that XYY tends to present with more severe social impairment and repetitive behavior. These discrepancies in ASD diagnoses in our study relative to prior work may be attributed to different methodology. Our study utilized rigorous gold-standard methodology to assess and diagnose ASD rather than relying solely upon questionnaire reports (for example, those used in Ross et al., 2012 or Samango-Sprouse et al., 2013) to characterize the ASD phenotype. Differences in diagnostic rates may also be driven by the co-occurrence of other forms of psychopathology in our sample. For instance, an increased prevalence in externalizing behaviors or hyperactivity in XYY may overshadow social communication deficits related to ASD [75]. ASD-related features may therefore be better appreciated and more readily diagnosed in XXY/KS, where externalizing tendencies are typically less severe. Finally, a third plausible explanation is that different sets of raters assessed ASD symptomatology across the XXY/KS and XYY cohorts. However, while between-rater idiosyncrasies in diagnostic tendencies may exist, all raters were doctoral-level clinicians with research-reliable administration and rating of gold-standard ASD assessments, so this explanation is improbable. It should also be noted that although Tartaglia et al. (2017) found much lower rates of ASD (10%) in their sample of 20 XXY/KS individuals, another study with a larger sample (n=51) documented ASD rates comparable to those in our study (27%, [53]) using the same gold-standard diagnostic tools. A third caveat to our study is that the observed profile of psychopathology in XXY/KS from our analysis of dimensional measures (and subsequently, our comparison with XYY) is shaped by the control group against which scores were compared. In this initial report, as well as in Raznahan et al. (2023), we have intentionally focused on a control group of XY individuals who have been screened to verify the absence of prior psychiatric diagnoses. Although this method allows for more precise detection of mental health difficulties in SCA groups, it likely inflates the magnitude of our effect sizes. Therefore, an important goal for future work would be use of alternative control groups including unscreened XY individuals without a genetic diagnosis. Fourth, the observational and cross-sectional nature of our study design means we cannot test the causal basis or development stability of observed associations. Securing such insights will have to await accrual of large longitudinal and interventional datasets which are especially hard to assemble in relatively rare GDDs. Fifth, our comparative analyses detail differences between group-level features of XXY/KS and XYY, but the dimensional data we include highlight the profound degree of interindividual variation within each of the SCA groups studied. Thus, while there is value and utility in understanding the average profiles associated with any given GDD and how these profiles may differ between GDDs, it is also essential to pursue complementary research approaches that examine sources of interindividual variation within each GDD [76].

#### Conclusions

Notwithstanding the above limitations and caveats, our study provides an unprecedentedly deep phenotypic description of mental health outcomes in XXY/KS and contrasts these with equivalent data in XYY syndrome. This effort advances clinical understanding of SCAs as important medical conditions in their own right, and provides a window into the relative influence of X– and Y-chromosome dosage on human development. The methods presented in this paper provide an expanded analytic tool kit which can now be generalized to other GDDs. Pursuing this broader effort will be important for clarifying if and how different GDDs vary in their profiles of psychopathological risk and their mappings between psychopathology and day-to-day functional outcomes. The answers to these questions will shape our theoretical models for genetic mechanisms of risk in psychiatry and our progress along the envisaged path towards genetically-informed precision psychiatry.

## Data Availability

All data produced in the present study are available upon reasonable request to the authors.

## ACKNOWLEDGMENTS

This study was supported by the intramural research program of the National Institute of Mental Health (NIMH) (NIH Annual Report Number: ZIAMH002949; Protocol: 89-M– 0006; ClinicalTrials.gov Number: NCT00001246). We thank the patients and their families for participating in this study, as well as the Association for X and Y Chromosome Variations (https://genetic.org) for their assistance with recruitment. We also thank Audrey Thurm, Lisa Joseph, and Cristan Farmer for their roles in data collection.

